# Functional dependence of COVID-19 growth rate on lockdown conditions and rate of vaccination

**DOI:** 10.1101/2021.06.06.21258425

**Authors:** F. Mairanowski, D. Below

## Abstract

It is shown that derived from the solution of differential equations analytical model adequately describes development epidemics with changes in both lockdown conditions and the effective rate of mass vaccination of the population. As in previous studies, the control calculations are in good agreement with observations at all stages of epidemic growth. One of the two model coefficients is uniquely related to the lockdown efficiency parameter. We obtained an approximate correlation between this parameter and the main conditions of lockdown, in particular, physical distancing, reduction in social contacts and strictness of the mask regime.

The calculation of the incident over a seven-day period using the proposed model is in good agreement with the observational data. Analysis of both curves shows that a better agreement can be obtained by taking into account the lag time of the epidemic response of about 10 days.

From the reverse calculation a time-varying curve of the infection rate associated with the “new” virus strain under mutation conditions is obtained, which is qualitatively confirmed by the sequencing data.

Based on these studies, it is possible to conclude that the ASILV analytical model developed here can be used to reliably and promptly predict epidemic development under conditions of lockdown and mass vaccination without the use of numerical methods.

The functional relationships identified allow us to conduct a rapid analysis of the impact of each of the model parameters on the overall process of the epidemic.

In contrast to previous studies, the calculations of the proposed model were performed using EXCEL, rather than a standard calculator. This is due to the need to account for multiple changes in lockdown conditions and vaccination rates.

## Introduction

Most models used to calculate the epidemic offer only numerical methods for solving. We developed a simple analytical model [1,2] that was used to analyse the spread of the coronavirus epidemic and showed that it adequately described the growth of infection under lockdown conditions. The transition from the absolute number of infected individuals to their relative number per inhabitant provided universal calculation ratios [3, 4, 5].

Performed control calculations, in which only one empirical factor was used, showed high accuracy of the results. In total, more than 30 test calculations have been carried out for settlements of widely varying population sizes - from individual urban areas in Berlin to major cities such as New York and several countries such as the United Kingdom, South Africa and Germany. The calculated curves are in good agreement with the corresponding statistical data. The correlation coefficients between the corresponding calculated and statistical curves reach values between 0.94 and 0.99.

The model was further developed to take into account the effects of abrupt changes in lockdown conditions and mass vaccination of the population [6]. A comparison of the results of calculations using this modified model with statistical observations for Israel shows good agreement. Three dimensionless complexes, made up of the intensities of the main processes: transmission, vaccination and lockdown restrictions, are found to determine the development of the epidemic.

The model was used to perform control calculations under different variants of changing lockdown conditions and vaccination rates. The analytical model, using functional relationships between the main parameters determining the epidemic development, makes it possible to assess the effectiveness of limiting the development of the epidemic by both lockdown and vaccination. Mass vaccination of the population is the most radical way of limiting the growth of the epidemic, whereas the introduction of a lockdown cannot prevent the development of an epidemic due to the high probability of new waves associated with the mutation of the virus.

Although there has been some success with this simple analytical model, there is a need to validate it further under different epidemic conditions and to use the model as a basis for further research in order to refine the basic patterns of epidemic spread.

## Methodology

Let us write the initial differential equations of the epidemic model, taking into account the impact of lockdown and mass vaccination on the epidemic spread, as [6]:

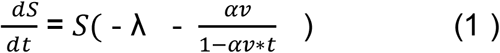

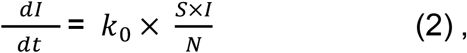

Where:

I - the number of infected persons at a given time,

*k*_0_ - coronavirus infection rate (1/day)

N - total population of the area under consideration,

S - the number of susceptible part of the population potentially capable of becoming infected due to contact with infected individuals.

λ - intensity factor of decrease in contacts of infected patients with persons who potentially can get infected by means of quarantine and other preventive measures.

v - population vaccination rate (1/day)

α - is the coefficient of vaccine effectiveness.

Equation (1), defines the change in the number of persons potentially susceptible to the virus under conditions of lockdown and mass vaccination of the population. The denominator in the last summand of equation (1) takes into account that as the proportion of the vaccinated population αv*t increases, the degree of impact of vaccination on the declining epidemic increases. The coefficient of effectiveness α depends on both the type of vaccine and the number of vaccination dose (first or second). We will assume that the maximum vaccination rate will not exceed (αv*t) _max_ ≤ 0.8, i.e. that with an 80% vaccination rate the epidemic cannot develop. This is a natural limitation of the proposed model. However, we have to take into account that some part of the population has already had the disease either explicitly or asymptomatically by the time mass vaccination begins.

The solution to equation (1) is as follows:

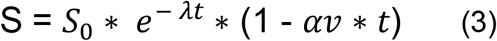

After substituting (3) into (2), solving the resulting equation, transformations and moving to a relative number of infections, we obtain the basic calculation equations.

For the period from outbreak to mass vaccination, *t* _*v*_ that is for t ≤ *t* _*v*_, when *αv* = 0, the solution of equation (2) has the form:

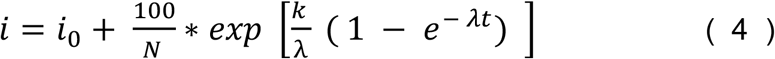

*i* - is the relative number of infected persons per one inhabitant of the settlement in question, as a percentage,

*i*_0_ - is the value of i at the initial moment of the calculation period,

K - is the transmission rate coefficient for the settlement with a population of N, which is calculated by the formula :

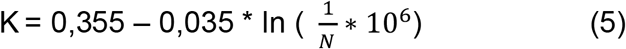

The K coefficient also depends on the transmissibility of the virus strain responsible for the epidemic spread during the period under consideration. The value of the first summand in (5) was obtained for the first and second waves of the virus epidemic. For further virus strains, we assume a higher value of 0.36. In the case where the spread of infection is associated with several virus strains, the calculated dependence will be written as follows:

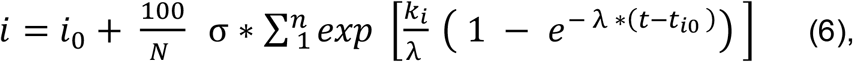

where:

i - is the sequence number of the strain of virus affecting the intensity of the epidemic over time *t* − *t*_*i*−1_,

*K*_*i*_ - the transmission rate coefficient of the new virus strain and the time of the epidemic wave associated with the new coronavirus strain

*t*_*i*0_ - the start time of the new epidemic wave associated with the new coronavirus strain.

σ - Heaviside symbol σ = 1 when t ≥ *t* _*i*_ и σ = 0 when t < *t*_*i*_

Dependence (6) is obtained under the assumption that the two or more virus species exist independently of each other.

Under conditions of mass vaccination when t ≥ *t* _*v*_, that is, when *αv* > 0:

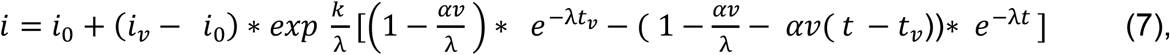

where *i* = *i*_*v*_ at t = *t*_*v*_

The calculations are performed first by (4) or (6) and then by (7) for the time period during which vaccination is carried out. The same equation (7) is used to calculate the spread of the epidemic under the condition of an abrupt change in vaccination rate, which was typical of many European countries, in particular Germany, for example.

In [5], an attempt is made to relate the model coefficient λ to the effectiveness of the lockdown condition. Let us make some specification of the relationship between this coefficient and the parameter L characterizing the level of reduction in the rate of growth of the epidemic due to lockdown

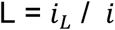

Where *i*_*L*_ and i are the intensity of the epidemic growth under lockdown and without lockdown, respectively. For example, if the application of lockdown reduces the maximum number of infected residents by half, then the coefficient L = ½ = 0.5. Using dependences (4) and (5) for time t → ∞ we find the relation between the coefficient λ and the parameter L. The graph of this dependence is shown in Fig. 1

**Figure 1.**
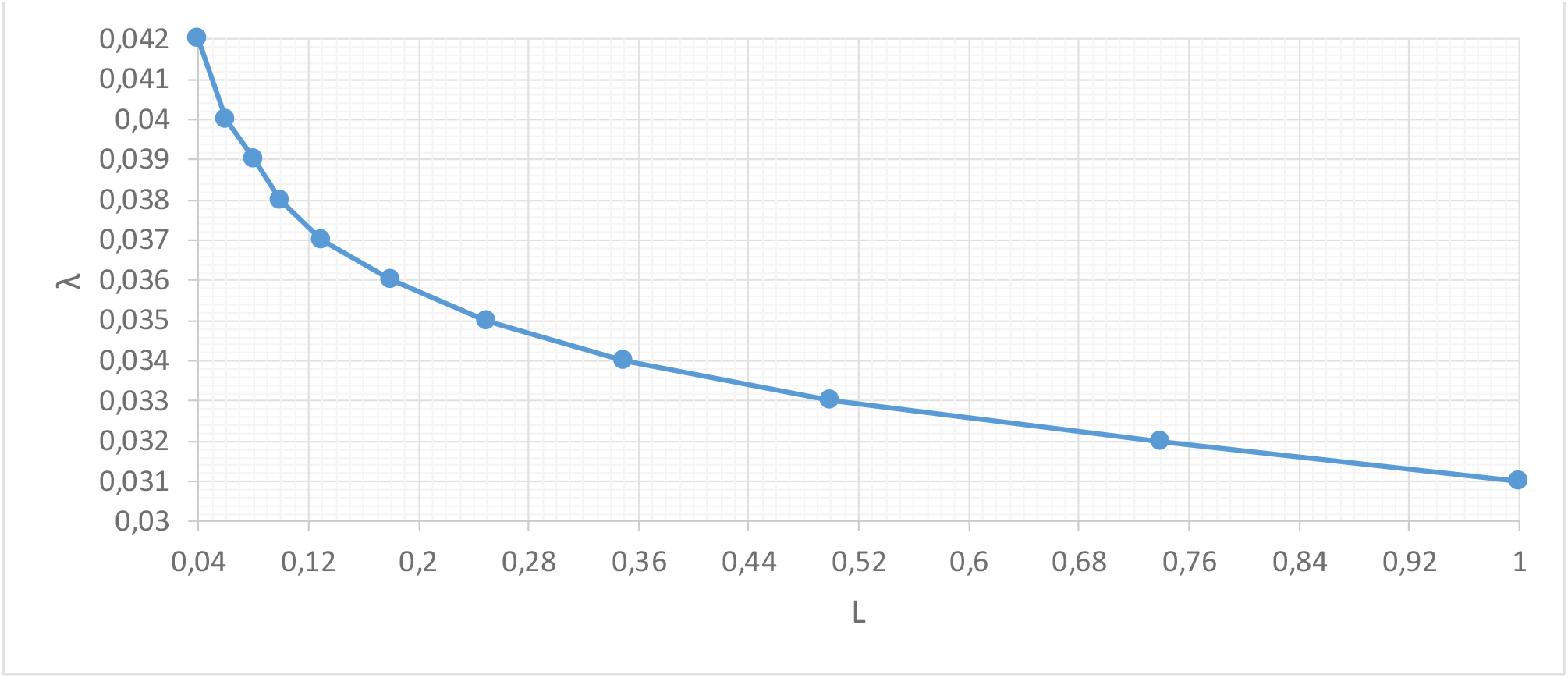
Dependence of the model coefficient λ on the lockdown efficiency L

This graph shows, in particular, that in the absence of lockdown, the coefficient λ can be assumed to be 0.031/day, and that when this coefficient is above 0.042 1/day, the epidemic wave is virtually suppressed by lockdown. However, this does not exclude the possibility of a new virus strain emerging when the lockdown conditions are relaxed. For the most typical values of λ = 0.034-0.035 1/day in most European countries, the L-factor varies between 0.2 and 0.3, i.e. lockdown can reduce the epidemic’s growth rate by a factor of 3-5.

The graph in Figure 1 can be approximated by the formula

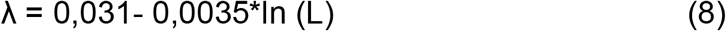

The relationship of both empirical coefficients of the model to population size, type of virus strain, lockdown conditions and population vaccination rate allows it to be used not only for analysing the development of an existing epidemic but also for operational forecasting of the development of COVID19. This model will hereinafter be abbreviated as ASILV (“analytical-susceptible-infection-lockdown-vaccination model”).

## Results

The proposed ASILV model was originally used to analyse the development of the epidemic and the impact of mass vaccination in Israel [6].

The results of the calculations and observations are in good agreement for both the pure lockdown and the lockdown with mass vaccination. As a second example of the applicability of the model, we will use it to perform a detailed analysis of the distribution patterns of the second and subsequent waves of the virus epidemic in Berlin. The analysis of various aspects of both the first wave and the further development of the epidemic in Berlin has been reviewed in our previous studies [1, 2, 3, 4, 5]. However, certain unresolved questions remain, related both to the analysis of the impact of abrupt changes in lockdown conditions, e.g. during the Christmas holidays, and to the role of mass vaccination of the population. For the first wave of the epidemic, a comparison of estimated and statistical data was presented in [4] and is shown in Fig. 2.

**Fig. 2.**
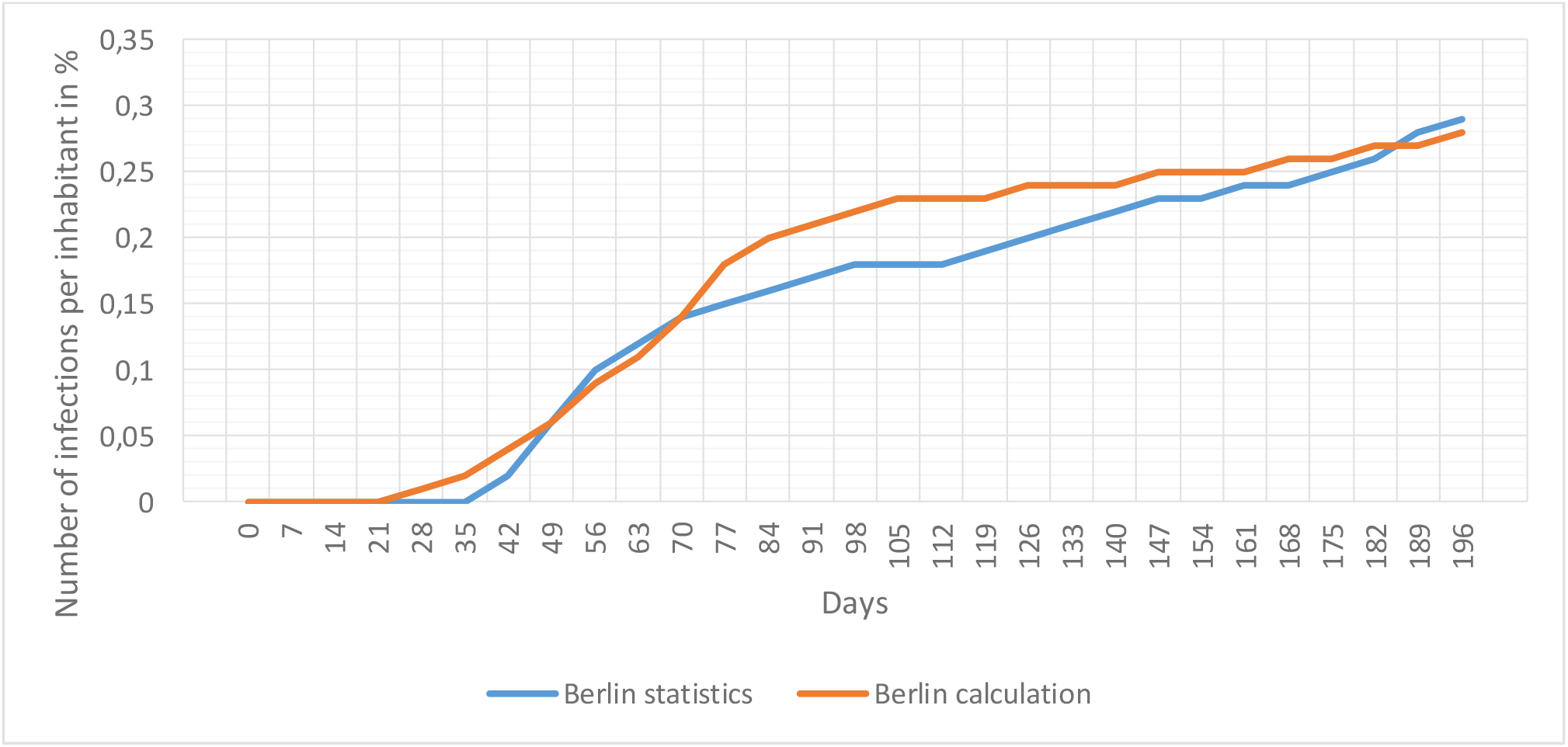
First wave of the COPID 19 epidemic in Berlin

Calculations were performed using relations (4) and (5) at a value of λ = 0.042 1/day. The start of the first wave was late February 2020; by 15 March, when the main lockdown measures were taken in Berlin, the total number of infected people did not exceed 300 and the daily increase in cases was about 20 people per day. This is why such a high coefficient λ was assumed in the calculations. In comparison, [4] in New York, for example, the lockdown was not imposed until more than 2 months after the epidemic had begun, when the total number of infected people had reached 37000, and the daily increase in cases was over 5000 per day. Accordingly, the coefficient λ at approximately the same level of lockdown was significantly lower, equal to 0.0345 1/day. Accordingly, while the total number of infected people in Berlin at the end of the first wave did not exceed 0.35% of the city’s population, in New York it reached 3%. The speed of response to the onset of an epidemic is thus one of the most important factors in controlling the spread of the infection. When analysing this graph, however, it must be taken into account that at the time of the first wave of the epidemic a sufficiently reliable service to determine the number of infected citizens had not yet been set up; some of the quantitative statistics are therefore not reliable enough. However, qualitatively, there is no doubt that the timely introduction of a lockdown can almost entirely prevent an increase in the epidemic. The first wave of the epidemic in Germany was over by June 15 when all the city’s infrastructure was fully operational. The calculations take 04.09.2020 as the start of the new epidemic wave in Berlin, when a monotonous increase in the number of infected people has been recorded [7]. The Berlin Senate adopted some restrictions for city services, sports and educational institutions from October 2020, but only on 9 November, i.e. 2 months after the beginning of the second wave of the epidemic, they were completely closed [8]. By this time, the total number of infected people in the city had reached 40,000 and the average daily increase in cases reached 1,000 per day [7].

Fig 3 shows the results of the statistics and calculations for the new virus wave from 04.09.2020.

**Figure 3.**
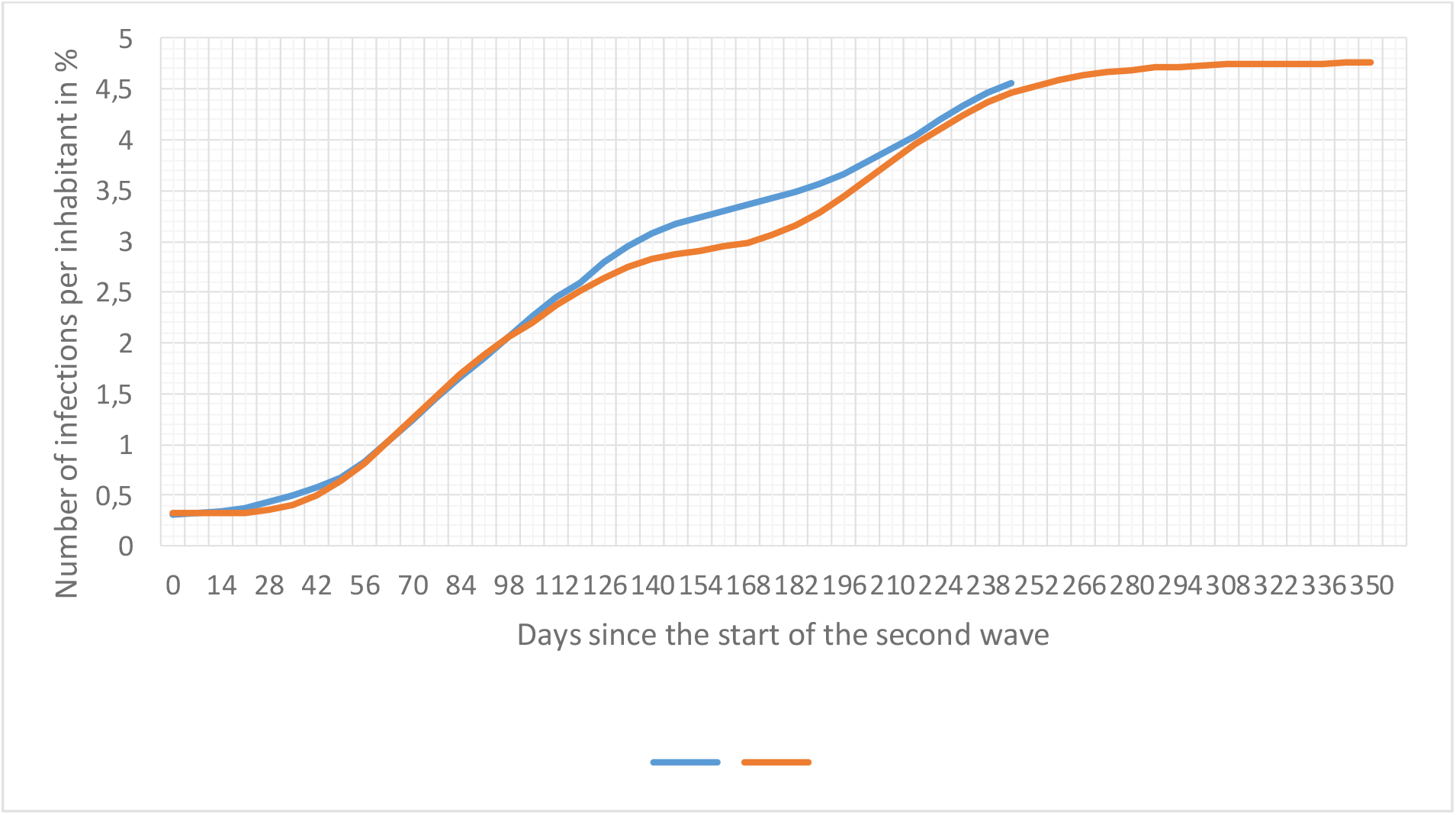
Development of the second and subsequent waves of the epidemic in Berlin. (i stat. - observed data, i calc.res - calculation for varying levels of lockdown, vaccination rate and climatic factors)

The calculations were performed with a time interval of 1 week (from Friday to Friday of the following week). During the first 98 days from the beginning of the second wave of the epidemic (i.e. until 18.12.2020), the lockdown conditions changed little. For this time period, as in our study [4], model coefficient λ = 0.035 1/day, and according to equation (5) K = 0.4 1/day.

Statistical data indicate a significant increase in the epidemic after about 100 days from the start of the second wave. Assuming that this period was characterized by weakening of the lockdown following Christmas and New Year’s celebrations, we performed the calculation according to dependence (7), conventionally assuming that lockdown restriction conditions were practically not met during the period 94 to 126 days (λ = 0.032 1/day). However, even such a rather exaggerated assumption about the breach of the lockdown did not allow us to obtain the calculated data satisfactorily agreeing with the observational data for the time period in question. This circumstance leads us to the most probable conclusion that the growth of the epidemic in this period is due to the emergence of a new strain of the virus. During the period from 126 to 140 days after the start of the second wave of the epidemic, the lockdown conditions should not have been disturbed. For that period of time (until about 22.01.2021), λ = 0.035 1/day was again assumed in the calculation model.

About 140 days after the beginning of the second wave of the epidemic in Berlin, there were noticeable signs of the emergence of a new so-called “British” strain of the B 1.1.7 virus. As virological studies have shown, this virus strain has a slightly higher transmissibility than previous strains [10]. In our model, this characteristic of the “British” strain was taken into account by increasing the K-factor. For subsequent calculations, this was assumed to be K= 0.41 1/day. Due to the sudden increase of epidemic intensity during this period in Berlin, additional restrictive lockdown measures were imposed. For this period, the tightening of lockdown conditions was taken into account in the calculations by increasing the coefficient λ to 0.036 1/day.

From the end of January 2021 onwards, a mass vaccination will be carried out in Berlin. However, the rate of vaccination in its initial phase was extremely low [9]. In the period from 22/01-16/04, i.e. 90 days, the percentage of Berliners vaccinated with the first dose of vaccine was 16% or less, with the second dose occupying about 7.5%. In the first stage it was mainly elderly citizens above 80 years of age who were relatively poorly involved in the transmission of the virus. Taking into account the effectiveness of vaccines in preventing transmission, we obtain a relationship for calculating the effective vaccination rate:

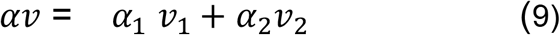

Vaccination rates for each vaccine dose *v*_1_ and *v*_2_ were calculated based on the data given in [9] as the ratio of the percentage of vaccinated population to the total time of mass vaccination of the population. The BionTech-Pfizer and Moderna vaccine efficacy ratios for the first and full dose of vaccination were taken as *α*_1_ = 0.7, *α*_2_ = 0.92 respectively [11].

In the second stage of vaccination (starting 16.04.2021, i.e. at t = 224), its intensity increased dramatically. By the time of writing this paper (02.06.2021) the vaccination rate of the first dose reached more than 43% of the city population, the second dose reached more than 19%.

Accordingly, the effective vaccination rate for the first period is αv = 0.0019 1/day, for the second αv = 0.0055 1/day.

As previously noted in [6], the development of an epidemic can be influenced by climatic factors. The dependence of the K coefficient on temperature and UV can be approximated using the following relationship

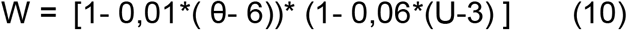

where W is coefficient of influence of climatic parameters on intensity of epidemic development, θ is average air temperature, U is value of UV index (for average conditions of Berlin it is assumed that θ = 6^0^ C, U=3). Accordingly, in addition to equation (5), we have:

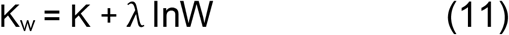

where Kw is the coefficient in equations (4), (6) and (7), taking into account the influence of climatic factors on it.

The calculation of this equation has revealed that starting from the second half of May, at the average maximum Berlin temperature of 25°C and the UV value of approximately 5 units the K coefficient is 0.39 1/day. Nevertheless, in view of the sharp decrease of the epidemic intensity at that time as a result of mass vaccination and the short duration of the period, this correction was not significant (about 10%). Nevertheless, these corrections were taken into account in the calculations starting from time t = 245 (07.05.2021).

The resulting calculated curve in Fig. 3 takes into account all changes in lockdown conditions and vaccination rates during the course of the epidemic, as well as changes in climatic factors affecting the intensity of the epidemic.

## Discussion

The results of calculations on the proposed ASILV model are in satisfactory agreement with the statistics data, both for the first epidemic wave (Fig.2) and for subsequent waves. The correlation coefficient between the calculated and statistical data for the second and subsequent waves is r = 0.9982. The proposed calculation methodology allows us to take into account, in a timely manner, the impact on the infection spread, both changes in the lockdown conditions, and the rate of vaccination.

Using the EXCELL software, it is also possible to quickly establish an incident rate over 7 days, one of the main characteristics determining the intensity of epidemic growth and which is adopted as the main criterion for mitigating lockdown, based on the ASILV model.

Fig. 4 compares the calculated and observed seven-day incident values for the second and subsequent epidemic waves (per 100,000 inhabitants). Overall, there is good agreement between the calculated and statistical data. The consistency of these results can be improved by shifting the estimated data to the right by about 10 days, i.e. taking into account the lagged response of the epidemic to the lockdown measure.

**Figure 4.**
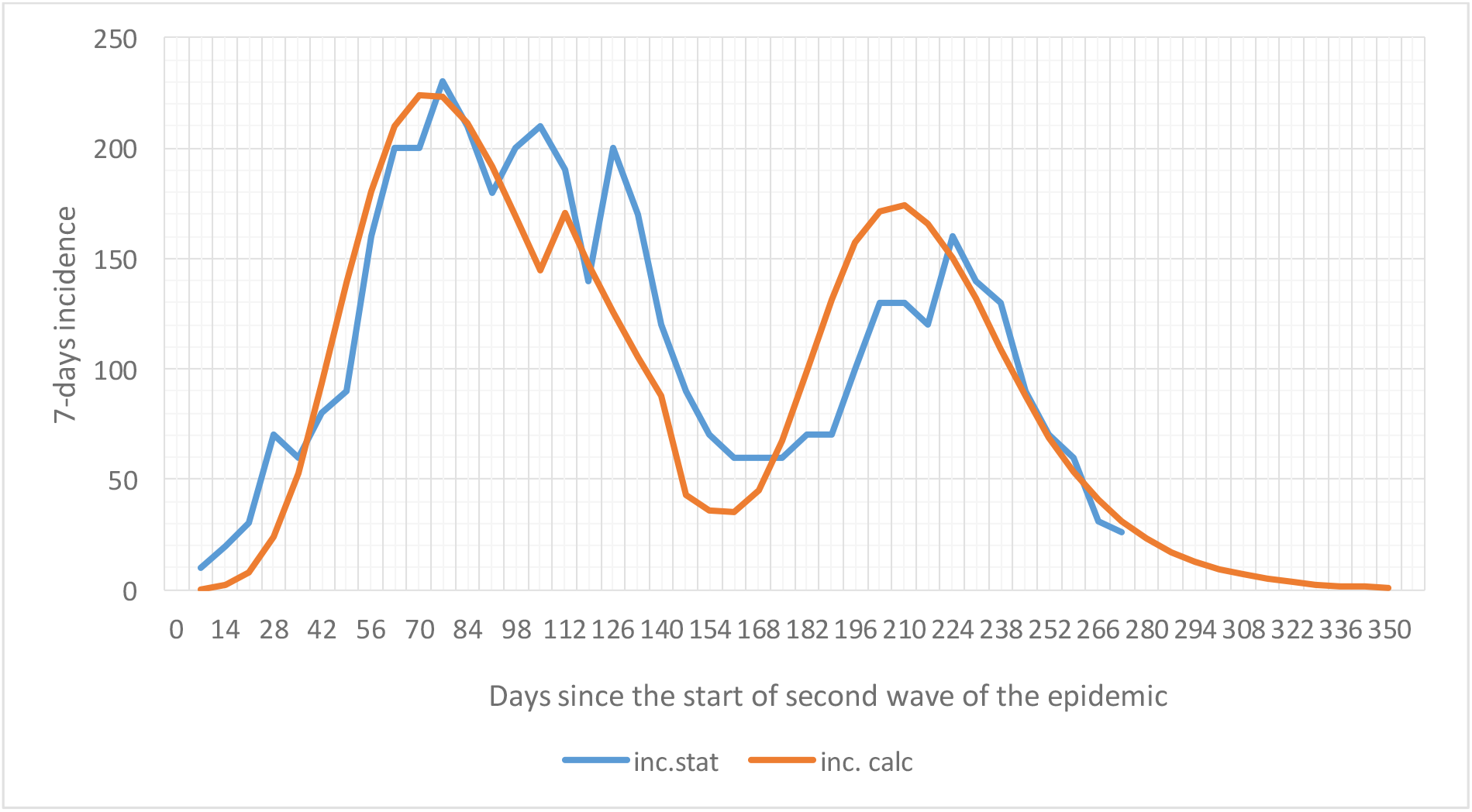
Incidence of epidemic growth over a seven-day period. (inc.stat. – observed data, inc.calc – calculation data)

Since abrupt changes in the epidemic growth rate are mainly due to a virus mutation and the emergence of a new strain that differs significantly from the previous one, the growth rate of the mutant virus variant was calculated in reverse.

The relative number of patients infected with the new virus strain (Fig. 5) was calculated as the ratio of the daily (weekly average) number of patients infected with the second virus strain s2 to the total daily increase in infections with coronavirus (s1+s2). These observations were obtained directly from virus sequencing [9,12]. For the calculations, the assumption was made that both virus strains acted independently. Overall, the qualitative agreement of the results is quite satisfactory. It should be noted that a much better agreement between the calculated and measured values of the growth intensity of the new strain could have been obtained by correcting for the lagged epidemic response, as can be seen in Fig. 4. It is important to emphasize that under conditions of mass vaccination the mutation of the virus can continue, but the development of the epidemic is halted. This is shown by the sequencing data, which indicate that the virus content determining the development of the epidemic does not exceed 92%.

**Fig. 5.**
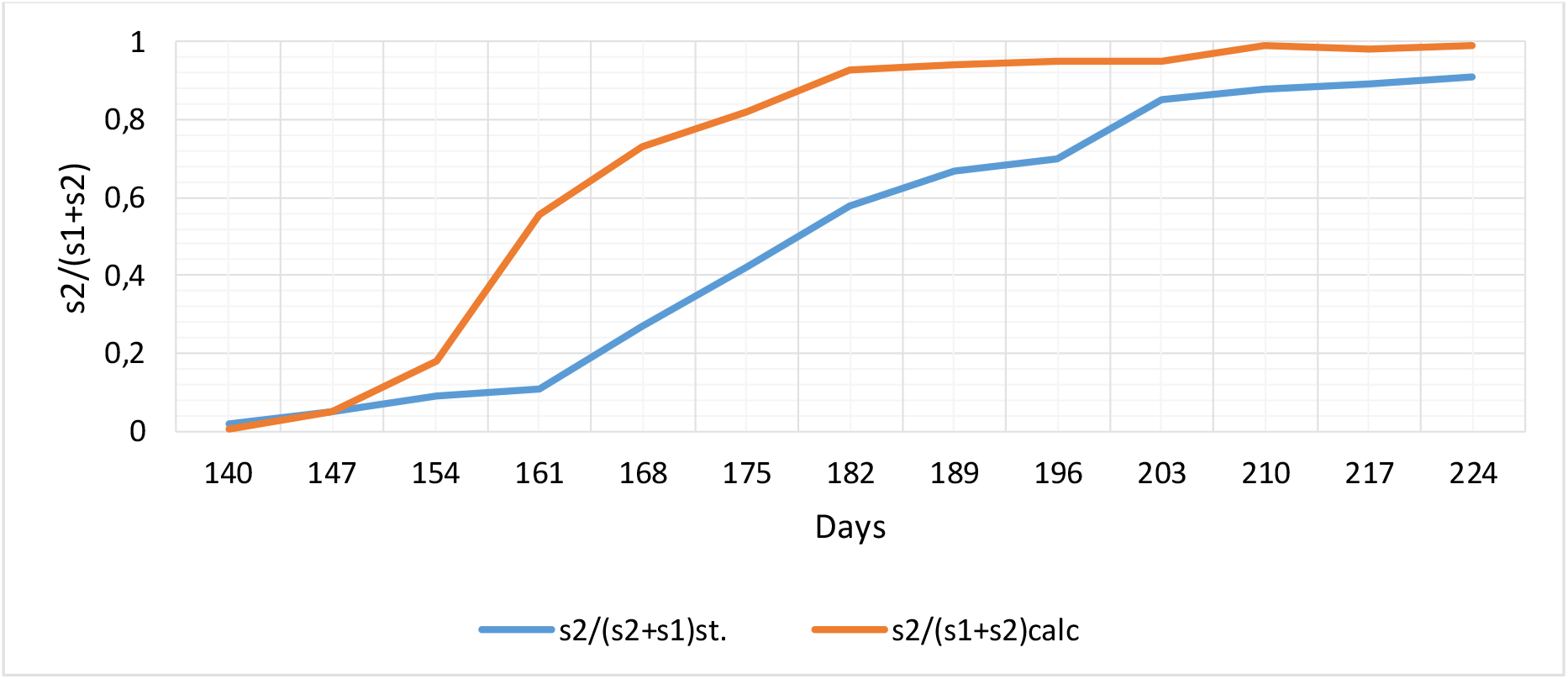
Intensity of growth of the new virus strain in Berlin.

At this stage of research it can be considered acceptable to use the assumption of independent action of both virus strains in model development.

For further development of the model and its use as a predictive model, a more detailed analysis of the dependence of the λ coefficient on the conditions of lockdown should be performed. The coefficient λ, as noted above, determines the effectiveness of reducing the epidemic growth rate L through the introduction of a lockdown, i.e. it depends on the conditions of its implementation. The main lockdown measures used to control the COVID-19 epidemic are: restriction of social contacts, compliance with the recommendation to maintain a distance of at least 1.5 metres between persons, wearing of protective masks in congested areas, compliance with hygienic standards, especially those related to hand washing and disinfection:

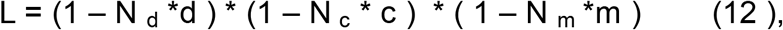

where:

N _d_, N _c_, N _m_ are the relative numbers of residents complying with the rule of physical distancing, reduction of social contacts and complying with the prescribed mask regime, respectively, d, c, m are the effectiveness of physical distancing, reduction of social contacts and mask mode, respectively, in limiting the spread of the epidemic.

The factors determining the effect of hand hygiene on the intensity of the epidemic are not included in this approximation. This is because the influence of hand hygiene can affect the epidemic in cases of lack of water for handwashing or lack of hygiene standards related to health education or tradition. In the Berlin context, as in Europe as a whole, hand hygiene problems are generally handled successfully. For example, UK members of the general population have a good level of understanding of the importance of adequate hand hygiene practice and compliance and its role regarding communicable disease prevention and control, according to surveys [13]. More than 90% of UK respondents consider hand hygiene to be important or very important for disease prevention (in comparison, about half as many respondents in the same survey consider vaccination to be important for disease prevention). About 10% of the population therefore do not perform the required hand hygiene, so even assuming that hand washing can reduce the COVIDS epidemic by 25% (this effect is achieved by hand disinfection) [14], even assuming this rate.

Maintaining a physical distance of 1 metre or more between individuals reduces the risk of transmission by 80% [15], i.e. d = 0.8. This risk of transmission is possible in the absence of a protective mask when the virus is transmitted together with aerosols. Cross-national surveys have estimated that about 75% of the German population would use a face mask all the time to prevent transmission (N _m_ = 0.75) [16]. Thus, it can be assumed that about 25% of the adult population, or N _d_ = 0.25, may be involved in transmission without protective masks.

Reduced social contacts also help to slow down the growth of the epidemic. According to studies [17], the average number of contacts in the United Kingdom during the epidemic was reduced by about 70% (c = 0.7). However, as approximately 60% of transmission is attributable to exposure to family members at home [17], the effective impact of social distancing on the epidemic is much lower, which suggests that N _c_= 1 - 0.6 = 0.4.

The final multiplier in formula (12) characterizes the effect of mask use on the growth rate of the epidemic. With regard to the efficacy of mask-based protection against infection, the m value depends on the type of mask used. In Germany, until mid-April, simple two-layer thin cotton fabric masks were the most common, with an average of m = 0.63 of particles [18]. Since mid-April a special decree of the Berlin Senate prohibited the use of these masks in transport, food shops and other public facilities. FFP 2 type masks, for which the aerosol blocking effect reaches m = 0.95 or higher, have been most widely used [19].

Let us make an approximate calculation of the value of the parameter L using the above approximate coefficients.

For two-layer fabric masks and partially FFP 2 type masks (our estimate for Berlin was about 15% of all protective masks) (m = 0.63* 0.85 + 0.94*0.15 = 0.7) L = 0.27. Thus, we find from relation (8) or from the graph in Fig.1 that for these conditions λ = 0.035 1/day. This value of λ has been taken for the calculations of virus spread in Berlin, both for the first and the second waves of epidemics. Under the conditions of widespread use of FFP 2 facemasks, the calculation according to formula (10) gives) L = 0.165 and correspondingly λ = 0.036 1/day. This is the value of the coefficient used in the calculations for the new epidemic wave in Berlin.

Particularly noteworthy is the effect of responsiveness on the onset of infection waves: it has been shown in many studies (e.g. [17]) that a delay in the introduction of a lockdown allows the virus to rapidly increase the number of infected individuals, so that even stricter lockdown measures subsequently fail to quickly quell the intense growth of the epidemic. This growth pattern was observed in the first wave of the epidemic, particularly in the United States (e.g. New York), Italy and Spain. The intense spread of the virus at the initial stage of the epidemic was fuelled by a lack of protective masks in a number of countries. In some countries (e.g. the United Kingdom, the United States, Spain, Sweden, etc.), national authorities failed to seriously assess the risks of epidemic spread and in virtually all countries there was a considerable delay in introducing a lockdown during the initial spread of the second and subsequent epidemics. The ratio (12) derived from the various countries’ statistics indirectly accounts for the impact of this delay on the growth of the epidemic. In the case of a rapid response to a situation of increasing infection, the coefficient λ should be increased, which was done, for example, in the calculation of the first wave of the epidemic in Berlin. It is difficult to obtain a quantitative relationship between delay time and efficacy in reducing the epidemic, both because in most countries the conditions for introducing lockdown have been continually adjusted, and because of the many breaches of lockdown. The best known violations are those that were partially responsible for the first wave of the epidemic, e.g. in France following a meeting of the Christian Open Door Church between 17 and 24 February in Mulhouse, which was attended by about 2,500 people, at least half of whom were believed to have contracted the virus [20] or as a result of a mass demonstration on 8 March in Spain [21]. The great diversity in response times and lockdown conditions during the first wave of the epidemic makes it difficult to systematize the relationship between λ and the magnitude of the delayed lockdown at the start of the epidemic. However, it can be estimated that if the response to the outbreak is rapid (within 2 weeks) and the lockdown rules are strictly observed, the λ increases by 20% compared to the result obtained from ratio (8). Thus, for example, if λ = 0.035 1/day is assumed to be the most realistic value for the second wave of an epidemic, then a rapid response to the start of an epidemic under similar lockdown conditions would increase the coefficient to 0.042 1/day. In this case, the introduction of a lockdown allows an epidemic to be quickly extinguished, as was the case, for example, during the first wave in Berlin. However, it should be borne in mind that a rapid exit from lockdown can lead to further waves of the epidemic due to mutations in the virus. If we assume an average of 5 mutations of the virus per year and the mutation probabilities are described by a Poisson distribution [6], we estimate the probability of one mutation occurring within 3 months to be around 70%.

During the second and subsequent epidemic waves, lockdown conditions and their implementation were almost identical in most countries, allowing ratios (12) and (8) to be used as estimates for determining the λ coefficient when using the ASILV model.

## Conclusions

1. The proposed analytical model ASILV adequately describes the development of the epidemic under various lockdown conditions and under variations in the rate of mass vaccination of the population. As in previous studies, the control calculations are in good agreement with observations at all stages of the epidemic’s growth.
2. One of the two model coefficients is uniquely associated with the lockdown efficiency parameter. The approximate correlation between this parameter and the main conditions of lockdown was obtained, in particular, physical distancing, reduction in social contacts, and the strictness of the mask regime, indicating that prompt introduction of restrictive measures is especially important in controlling an epidemic, both at the beginning of infection and when a new strain of the virus emerges.
3. The seven-day incident calculations of the ASILV model are in good agreement with observations. Analysis of both curves suggests that a better agreement can be achieved if the time lag of the epidemic response of about 10 days is taken into account in the calculations.
4. The time-varying curve of the relative impact of the “new” virus strain under mutation conditions obtained from the back-calculation is qualitatively confirmed by the observations.
5. It is possible to conclude from the fulfilled research to use the developed analytical model ASILV for the forecasting of the epidemic development in conditions of lockdown and mass vaccination. One of the model coefficients is functionally related to lockdown conditions, the other one to population number and, to a smaller extent, to virus transmissibility and climatic factors.
6. The functional relationships found allow the impact of each of the model parameters on the overall process of development of the KOVID-19 epidemic to be analysed operationally.

## Data Availability

all data referred to in the manuscript and note links below is regard

https://www.instagram.com/felma39/

## Reference

[1] Below, D., & Mairanowski, F. (2020). Prediction of the coronavirus epidemic prevalence in quarantine conditions based on an approximate calculation model. medRxiv.

[2] Below, D., Mairanowski, J., & Mairanowski, F. (2020). Checking the calculation model for the coronavirus epidemic in Berlin. The first steps towards predicting the spread of the epidemic. medRxiv.

[3] Below, D., Mairanowski, J., & Mairanowski, F. (2021). Analysis of the intensity of the COVID-19 epidemic in Berlin. Torwards an universal prognostic relationship. medRxiv.

[4] Below, D., Mairanowski, J., Mairanowski F. (2021) Comparative analysis of the spread of the COVID 19 epidemic in Berlin and New York City based on a computational model. https://www.medwinpublishers.com/PHOA

[5] Below, D., Mairanowski, J., Mairanowski F. (2021). Development of the COVID19 epidemic model: calculations for a mutating virus. https://medwinpublishers.com/JQHE/

[6] Below, D., Mairanowski F. (2021). The impact of vaccination on the spread patterns of the COVID epidemic. medRxiv.

[7] Development of number of Coronavirus cases: Berlin, Germany. https://coronalevel.com/Germany/Berlin/

[8] Der Regierende Burgomeister von Berlin-Senatskanzlei. Archiv der Änderungsverordnungen zu Infektionsschutz-und Eindämmungsmaßnahmen - Berlin.de

[9] Der Regierende Burgomeister von Berlin-Senatskanzlei. Corona-Lagebericht - Berlin.de

[10] Nicholas G. Davies, Sam Abbott, et all. Estimated transmissibility and impact of SARS-CoV-2 lineage B.1.1.7 in England. Science 03 Mar 2021: https://science.sciencemag.org/content/early/2021/03/03/science.abg3055

[11] Noa Dagan, M.D., Noam Barda, M.D., Eldad Kepten, Ph.D., Oren Miron, M.A., Shay Perchik, M.A., Mark A. Katz, M.D., Miguel A. Hernán, M.D., Marc Lipsitch, D. Phil. BNT162b2 mRNA Covid-19 Vaccine in a Nationwide Mass Vaccination Setting. https://www.nejm.org/doi/full/10.1056/NEJMoa2101765

[12] RKI. Bericht zu Virusvarianten von SARS-CoV-2 in Deutschland. Stand 19.05 2021.https://www.rki.de/DE/Content/InfAZ/N/Neuartiges_Coronavirus/DESH/Bericht_VOC_2021-05

[13] Aaron Lawson et.al. An Investigation of the General Population’s Self-Reported Hand Hygiene Behaviour and Compliance in a Cross-European Setting. Int. J. Environ. Res. Public Health 2021, 18, 2402. https://doi.org/10.3390/ijerph18052402)

[14] Wang Y, Tian H, Zhang L, et al. Reduction of secondary transmission of SARS-CoV-2 in households by face mask use, disinfection and social distancing: a cohort study in Beijing, China. BMJ Global Health. 2020; doi:10.1136/bmjgh-2020-002794

[15] Derek K Chu, et al. Physical distancing, face masks, and eye protection to prevent person-to-person transmission of SARS-CoV-2 and COVID-19: a systematic review and meta-analysis. Lancet 2020; 395: 1973–87. Published Online June 1, 2020 https://doi.org/10.1016/S0140-6736(20)31142-9

[16] Wearing face masks outside during the coronavirus pandemic in Europe 2021, by country. Published by Conor Stewart, Jan 12, 2021. https://www.statista.com/statistics/1114375/wearing-a-face-mask-outside-in-european-countries/

[17] Jarvis et al. Quantifying the impact of physical distance measures on the transmission of COVID-19 in the UK. BMC Medicine (2020) 18:124 https://doi.org/10.1186/s12916-020-01597-8

[18] Eugenia O’Kelly et al. How well do face masks protect the wearer compared to public perceptions? medRxiv preprint doi: https://doi.org/10.1101/2021.01.27.21250645

[19] D. Lepelletier et al. What face mask for what use in the context of the COVID- 19 pandemic? Journal of Hospital Infection 105 (2020) https://www.journalofhospitalinfection.com/article/S0195-6701(20)30211-5/fulltext

[20] Coronavirus : la “bombe atomique “du rassemblement évangélique de Mulhouse”. Le Point. 28 March 2020.

[21] Spain goes on nationwide lockdown as coronavirus cases surge. Washington Post. 15 March 2020. Retrieved 15 March 2020.

